# Prediction of chronological age from resting-state EEG power in the first three years of life

**DOI:** 10.1101/2024.05.31.24308275

**Authors:** Winko W. An, Aprotim C. Bhowmik, Charles A. Nelson, Carol L. Wilkinson

## Abstract

The infant brain undergoes rapid and significant developmental changes in the first three years of life. Understanding these changes through the prediction of chronological age using neuroimaging data can provide insights into typical and atypical brain development. We utilized longitudinal resting-state EEG data from 457 typically developing infants, comprising 938 recordings, to develop age prediction models. The multilayer perceptron model demonstrated the highest accuracy with an R^2^ of 0.82 and a mean absolute error of 92.4 days. Aperiodic offset and periodic theta, alpha, and beta power were identified as key predictors of age via Shapley values. Application of the model to EEG data from infants later diagnosed with autism spectrum disorder or Down syndrome revealed significant underestimations of chronological age. This study establishes the feasibility of using EEG to assess brain maturation in early childhood and supports its potential as a clinical tool for early identification of alterations in brain development.

## 1 Introduction

Over the first three years of life, the infant brain undergoes dramatic changes across multiple scales. These developmental processes include structural increases in both cortical volume [1] and surface area [2], as well as changes occurring at the level of synaptogenesis [3, 4], inhibitory neuron migration and maturation, and network formation [5]. It is presumed that the coordinated timing of these foundational neurodevelopmental processes is generally similar across infants, and that significant alterations in these processes would impact later brain functioning and in turn cognition and behavior.

One way to study brain development across multiple scales is through the prediction of chronological age based on neuroimaging data [6]. The concept of ‘brain age’ largely began in the area of aging research, with the goal of better understanding differences between healthy brain aging versus advanced brain aging associated with neurodegenerative disorders [7, 8]. However, there is growing interest in similarly understanding the developing brain and how differences in predicted versus chronological brain age (termed the ‘brain age gap’) relate to various neurodevelopmental conditions such as autism spectrum disorder, attention deficit/hyperactivity disorder, and intellectual disability [9–11]. It is hypothesized that, if a child has altered early brain development, their estimated brain age will be substantially different from their chronological age (i.e., a more negative brain age gap). Thus far, the majority of brain age studies have focused on predicting across a broad age range, from infancy to adulthood, with a very limited number of samples in infants and toddlers [12, 13]. This limited study of early development prevents a fine-grained evaluation of brain milestones critical for developmental processes and reduces opportunities to develop clinical tools effective for early screening. In addition, no published studies have utilized EEG to predict age specifically in infants and toddlers, and only a handful have been published utilizing structural MRI and diffusion tensor imaging [14–16]. Finally, predictive brain age studies often do not include investigation of feature importance within machine learning models, preventing identification and characterization of features relevant to developmental brain maturation.

Using resting state longitudinal EEG data collected from 457 typically developing infants (938 EEG recordings) we first demonstrate the accuracy of four different linear or nonlinear machine learning models in predicting chronological age using resting state EEG features. Then, using an innovative approach that combines Shapley values and hierarchical clustering we identified clusters of features (aperiodic offset and periodic alpha and beta band power) contributing most to age prediction models. Finally, to explore whether developmental, EEG-based, brain age predictive models could be used as a marker of altered brain development we applied our model to EEG data collected from either infants and toddlers later diagnosed with autism spectrum disorder (ASD) or infants and toddlers with Down syndrome (DS). In the ASD group, our model significantly underestimated chronological age at the oldest age bin (1000 – 1150 days), but not at younger bins; in the DS group, this underestimation of age was significant across the full range of age prediction.

## 2 Results

### 2.1 Prediction of chronological age from EEG power

Resting-state EEG data collected in the same laboratory from typically developing infants and toddlers aged 2 – 38 months were used to create predictive models. A set of 18 features from each of 4 regions of interest (frontal, temporal, central, posterior) were calculated from aperiodic and periodic power spectra using an automated pipeline to enhance reproducibility. Absolute, aperiodic, and periodic power spectra by age bins are shown in Figure 1. Features included measures characterizing aperiodic and periodic activity and principal components in pre-defined frequency bands (See Methods for more details).

**Fig. 1.**
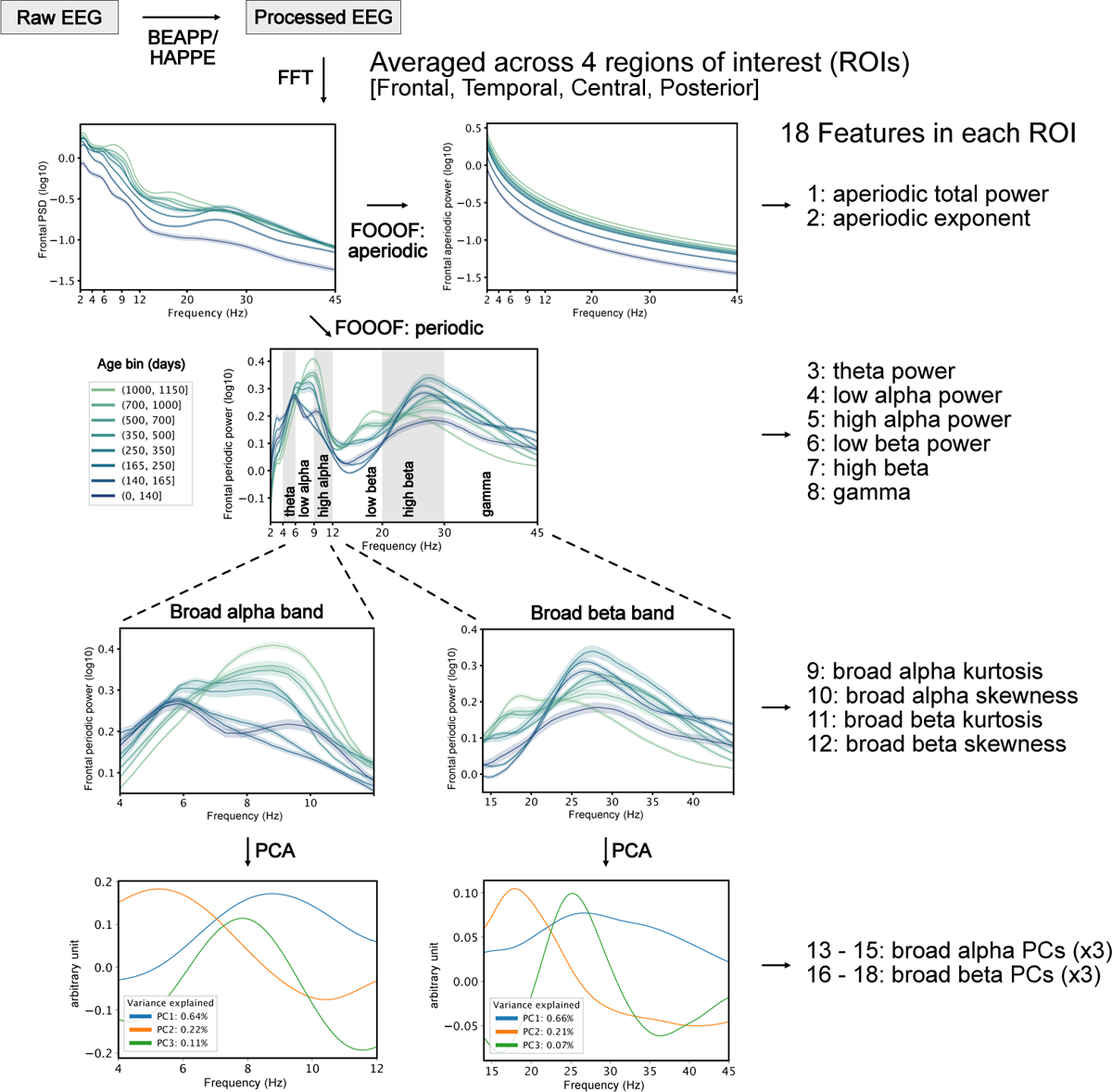
Feature extraction steps in this study. Raw EEG signals were cleaned using an automated processing pipeline: BEAPP [20] and HAPPE [21]. Power spectral density (PSD) was calculated from each electrode in processed EEG via Fast Fourier Transform (FFT) and averaged across four regions of interest (ROIs): frontal, temporal, central, and posterior. Then, the PSD was parametrized into aperiodic and periodic components using SpecParam [22]. From the aperiodic component, we extract two features in each ROI: total power and exponent. From the periodic component, first, we extracted six features of power, one for each canonical frequency band: theta (4 – 6 Hz), low alpha (6 – 9 Hz), high alpha (9 – 12 Hz), low beta (12 – 20 Hz), high beta (20 – 30 Hz), and gamma (30 – 45 Hz). Second, we analyzed the periodic component within the broad alpha (4 – 12) and broad beta (14 – 45) bands, as we observed a strong age effect on the shape of PSD in these bands. From each band, we extracted two features of power distribution: kurtosis and skewness. Additionally, we applied a principal component analysis (PCA) and extracted three features (PC1 – 3) in each band — together, they accounted for *>*90% of total variance in each band. In total, we extracted 72 features from each EEG sample — 18 features in each of the 4 ROIs.

With these 72 features we trained and tested four distinct regression models: Lasso, Random Forest, XGBoost, and Multilayer Perceptron (MLP). Detailed model specifications are provided in Methods, and the architecture of the MLP is shown in Figure 2B. Chronological age was reliably predicted from EEG in all four models (Figure 2C). The MLP yielded the best results among all models with an R^2^ of 0.82 and a mean absolute error (MAE) of 92.9 days. Notably, this MAE accounts for a mere 8.5% of the age range covered in our dataset, which spans from 59 days to 1150 days. The other three models had MAE ranging from 113.4 – 131.6 days, with R^2^ between 0.69 and 0.76 (Figure 2C). In all four models, the variance in prediction errors increased with age, suggesting the presence of heteroskedasticity in the regression analysis (Figure 2D), which was confirmed by a Breusch-Pagan Lagrange Multiplier test (p*<*0.001 in all models). This observation justified (and originally motivated) the application of the Yeo-Johnson transformation to chronological age prior to model training (for details, see Section 5.4). Additionally, the absolute prediction error significantly correlated with age, in which a positive error (i.e., predicted age *>* actual age) was associated with a younger age (Pearson’s r*>*0.034, p*<*0.001 in all models). Similar observations have been reported in previous brain age prediction studies, and is commonly referred to as “age bias” in prediction error [17–19].

**Fig. 2.**
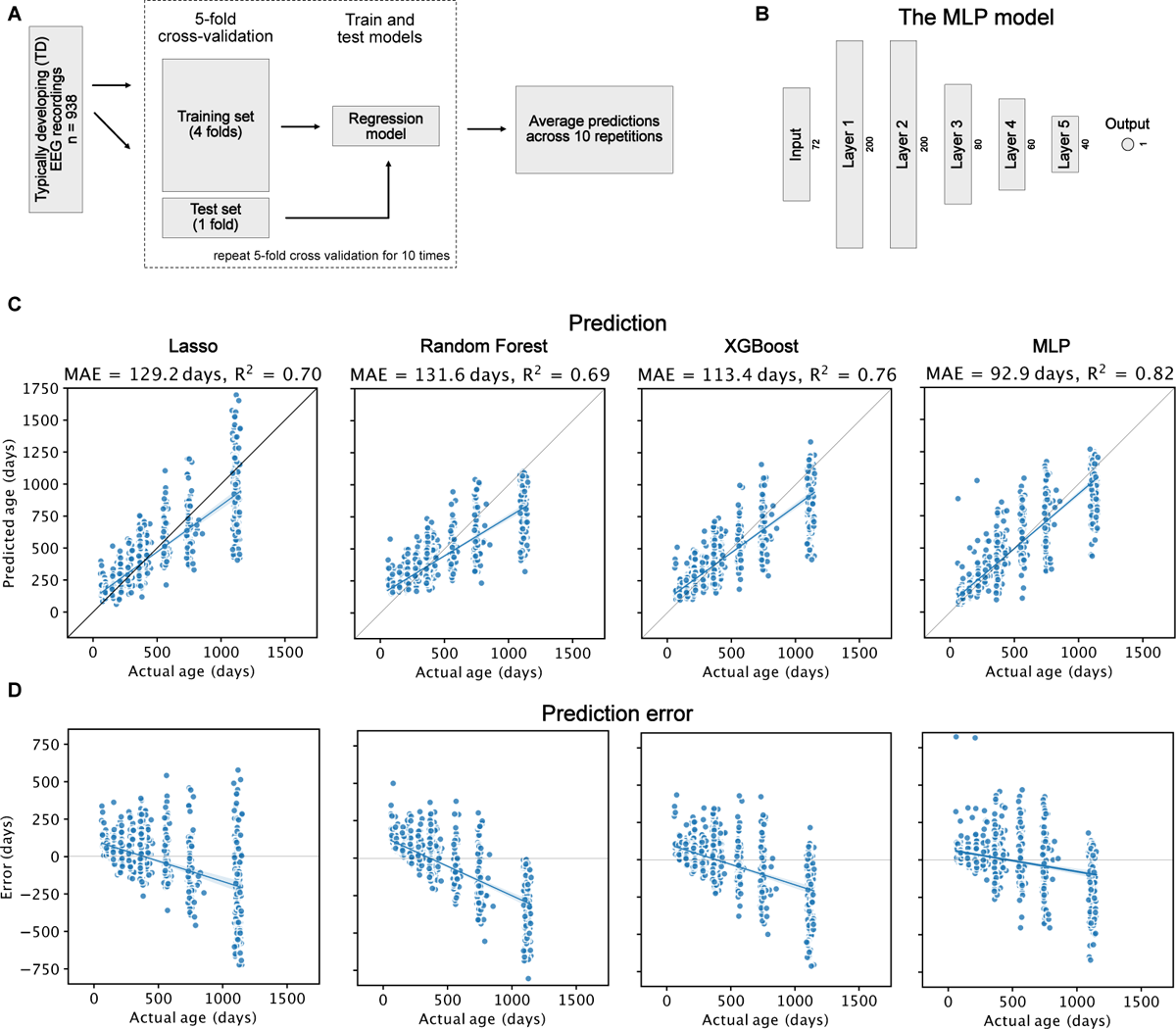
A: The schematic diagram of training and test steps. B: The architecture of the multi-layer perceptron (MLP) model in this analysis. C: Prediction results for each of the four regression models. The mean absolute error (MAE) and R^2^ of each model are shown in the headings. The diagonal represents perfect prediction (i.e., y = x). D: Prediction error for each of the four models.

### 2.2 Importance of EEG features in age prediction

Next, we analyzed feature importance to identify EEG features that drive model prediction and thus are tightly associated with early brain development. We employed the Shapley Additive Explanations (SHAP), a game theoretic approach to explain the output of machine learning models [23]. This algorithm assigns a value to *each feature* within every EEG sample (each recording), quantifying how much one feature shifts a model prediction from a baseline output (the average of all training samples). Feature importance can then be inferred from the absolute value of SHAP. Multicolinearity is a common challenge in EEG data that can cause misleading interpretations of feature importance [24]. To solve this problem, we leveraged the additive property of SHAP [25], where the joint importance of two features can be estimated by the sum of their SHAP values; we split the 72 features into 11 clusters (Figure 3C) using hierarchical clustering and summed SHAP values within each feature cluster (Cluster-SHAP). Cluster importance was then evaluated from the mean absolute value of Cluster-SHAP across samples (mean *|*Cluster-SHAP*|*) and the correlation between Cluster-SHAP and actual age. In both metrics, a higher value indicates greater cluster importance [26].

**Fig. 3.**
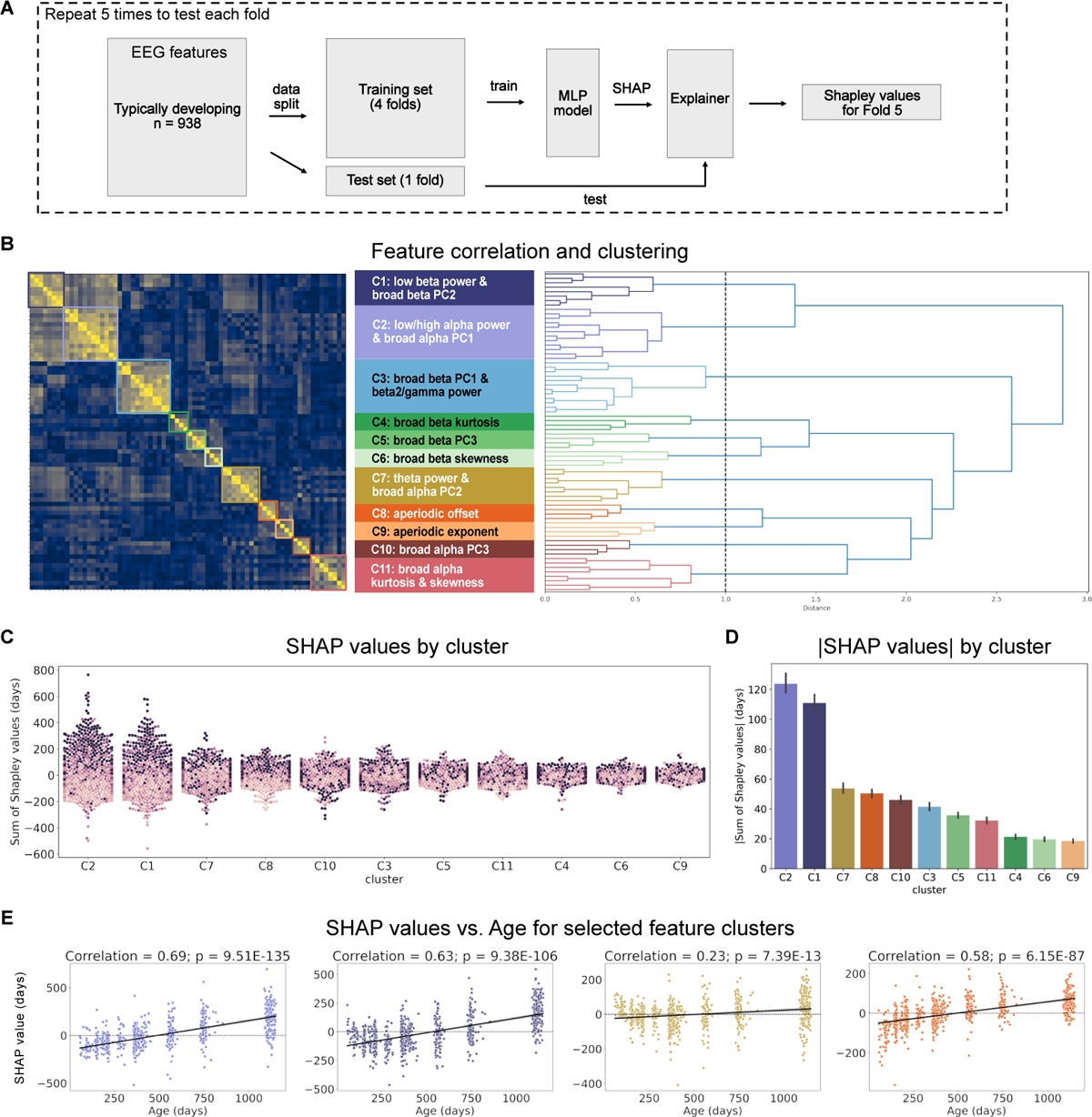
A: Schematic diagram for the feature importance analysis. B: Correlation between each pair of EEG features (left) and the hierarchical clustering (right) based on feature correlation. Each cluster contains different types of features and is labelled with a color; the same labelling scheme applies to all figures in this paper. C: SHAP values in each feature cluster. SHAP value was calculated for each feature in each sample (i.e., EEG recording). Every point in this figure represents the SHAP value summed across all features in a cluster in one sample; the darkness of color represents the age of this sample. D: The average absolute SHAP values across all samples in each feature cluster. Error bars indicate standard error. E: Relationship between SHAP values and age in four selected feature clusters. Black line represents the best linear fit. Correlation was calculated using Pearson’s r.

Using the above approach, we identified two clusters of features with the highest mean*|*Cluster-SHAP*|* — C2, consisting of features in the periodic alpha band (low/high alpha power and PC1 in broad alpha band), and C1, consisting of features in the periodic beta band (low beta power and PC2 in broad beta band) (Figure 3D; Table 1). They were followed by C7 and C8, consisting of features in the theta band and aperiodic offset, respectively. The Cluster-SHAP values in these feature clusters, especially in C1, C2, and C8, also significantly correlated with age (Pearsons’s r*≥*0.23, p*<*0.001; Figure 3E; Table 1). This further validates the importance of these features as they associated a more positive shift in model output with older chronological age, facilitating more accurate predictions. In summary, we identified features in aperiodic offset and periodic theta, alpha, and low beta band as important drivers of age prediction.

**Table 1.** The absolute values of Cluster-SHAP and their correlation with age in each feature cluster. Rows are sorted from the highest *|*Cluster-SHAP*|* to the lowest.

| Cluster | Features | \|Cluster-SHAP\|<br>mean (std) | Correlation with age |  |
| --- | --- | --- | --- | --- |
|  |  |  | r | p |
| C2 | low/high alpha power,<br>broad alpha PC1 | 120.22 (94.07) | 0.69 | <0.001 |
| C1 | low beta power,<br>broad beta PC2 | 110.66 (87.06) | 0.63 | <0.001 |
| C7 | theta power,<br>broad alpha PC2 | 54.10 (50.40) | 0.23 | <0.001 |
| C8 | aperiodic offset | 50.29 (42.80) | 0.58 | <0.001 |
| C10 | broad alpha PC3 | 46.69 (46.54) | 0.02 | 0.517 |
| C3 | broad beta PC1,<br>high beta/gamma power | 42.43 (42.07) | 0.07 | 0.031 |
| C11 | broad alpha kurtosis<br>and skewness | 36.87 (35.77) | 0.21 | <0.001 |
| C5 | broad beta PC3 | 35.25 (29.99) | 0.25 | <0.001 |
| C6 | broad beta skewness | 20.28 (20.87) | 0.12 | <0.001 |
| C4 | broad beta kurtosis | 19.56 (22.07) | 0.10 | 0.002 |
| C9 | aperiodic exponent | 18.87 (18.64) | 0.16 | <0.001 |

### 2.3 Prediction of age in children with autism or Down syndrome

To assess whether an EEG-based brain-age prediction model in early development could be useful in early identification of children with neurodevelopmental disorders, we applied our model on EEG data from infants later diagnosed with ASD or with Down syndrome (DS) and compared its prediction with that of the TD group. To assess prediction accuracy within the ASD cohort, we first created a test set with data from children with ASD and age-matched TD peers (n = 246 EEGs in each group). We divided the remaining 692 TD samples into 10 folds with equal sizes and trained our best-performing MLP model (Figure 2B) on 9 random folds. We repeated this process 10 times, each time with a different fold left-out for testing, and the outputs from these 10 models were averaged as the final prediction for each test sample. As expected, age of the TD children was accurately predicted by this model (MAE = 100.2 days); the fitted prediction line for the TD group almost overlapped with the line of perfect prediction (Figure 4A). For ASD infants, prediction was comparatively less accurate (MAE = 107.2 days) with age being underestimated, especially at older ages. A series of two-sample t-tests were conducted to compare the prediction errors between groups at pre-defined age bins (Table 3). A significant difference was only observed at the oldest age bin (1000 – 1150 days; p = 0.007, Cohen’s d = 0.87; Figure 4B). We then conducted an exploratory analysis to compare the EEG features between groups within this oldest age bin via two-sample t-tests. In the frontal ROI, for example, significant group differences were observed in broad beta band skewness (p = 0.003), broad alpha band skewness (p = 0.022), and aperiodic exponent (p = 0.017). Four other features (theta power, aperiodic offset, broad alpha band PC3, and broad beta band PC3) showed trending significance in their difference between the two groups (0.05*<*p*<*0.1).

**Fig. 4.**
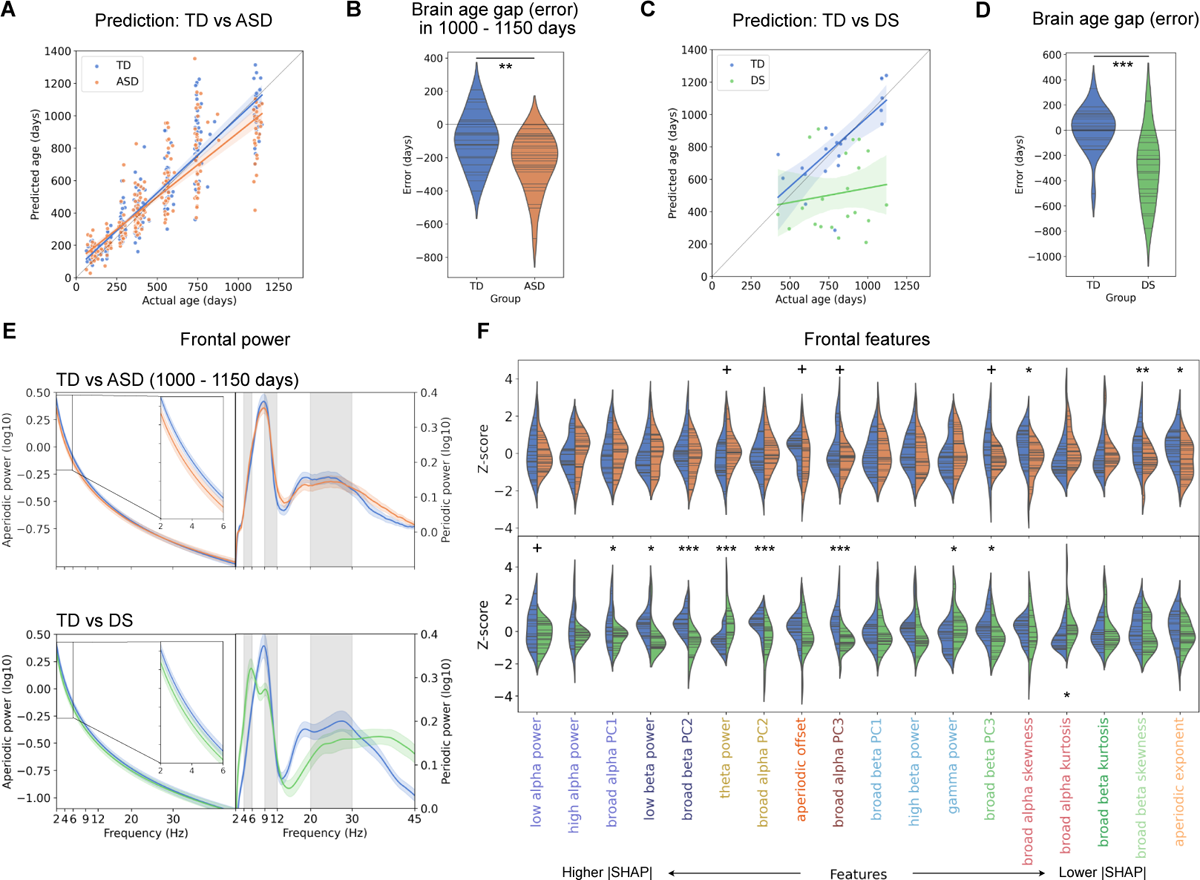
Comparing brain age prediction between children with typical development (TD) and neurodevelopmental disorders. A: Age prediction in TD and ASD using a MLP model depicted in Figure 2B. B: Brain age gap comparison between ASD and TD in 1000 – 1150 days. C: Age prediction in TD and DS using the same MLP model. D: Brain age gap comparison between DS and TD. E: Aperiodic and periodic power in the frontal ROI in TD vs ASD in 1000 – 1150 days (upper) and TD vs DS (lower). F: Exploratory feature comparisons, TD vs ASD in 1000 – 1150 days (upper) and TD vs DS (lower), in the frontal ROI as an example. Each feature was denoted as a Z score after being normalized by the mean and standard deviation across two groups. +, p*<*0.1; *, p*<*0.05; **, p*<*0.01; ***, p*<*0.001

A similar set of analyses were conducted to compare age prediction between TD and DS groups, where DS (n = 23) and age-matched TD samples were reserved for testing, and the remaining TD samples were used to train the MLP model. Prediction for the TD group (MAE = 107.0 days) was more accurate than that for the DS group (MAE = 331.9 days), in which the model substantially underestimated the age of children with DS (Figure 4C & 4D); brain age gap was significantly more negative in the DS than in the TD group (p*<*0.001, Cohen’s d = 1.32). Different from the comparisons between TD and ASD, more features with high importance showed significant difference between TD and DS (Figure 4F).

## 3 Discussion

Here we demonstrate that resting-state EEG features, extracted from aperiodic and periodic power spectra, can reasonably predict chronological age early in child development via a multi-layer perceptron (MLP) model. With an innovative approach combining Shapley values and hierarchical clustering, we identified two important clusters of features, namely the periodic alpha and beta band power, that contributed most to age prediction. In addition, as a proof of concept, we tested this MLP model’s ability to capture early signs of neurodevelopmental conditions by applying this model to EEG collected from children who later received an ASD diagnosis or children with DS. The model significantly underestimated chronological age in toddlers with ASD and in infants and toddlers with DS, supporting the brain age gap as a potential tool for early identification of delays or alteration in early brain development.

Our work is one of the first studies investigating EEG-based age prediction across the first three years of life; only a handful of published neuroimaging-based age-prediction studies overlap with our age range (Table 2), and our MLP model yielded comparable results. Built upon one of the most extensive neuroimaging samples of infants and toddlers to date, our study exclusively focuses on early brain development. A model dedicated to the infant-toddler period facilitates the detection of the rapid developmental changes occurring both structurally and functionally during this period [27]. In doing so, it also improves the model’s potential in identifying early indicators of alterations in brain development. While surprisingly few studies have used EEG in brain-age prediction, EEG may be an ideal candidate for models focused on infant and toddler years. Unlike MRI, EEG is low-cost and can be collected in awake and moving children, allowing for frequent data collection without sedation. In addition, EEG is a direct measurement of neuronal activity reflective of underlying network circuitry and therefore can capture the development and maturation of functional networks.

**Table 2.** Model performance comparison with literature MAE: mean absolute error; sMRI: structural MRI.

| Study | Modality | Age range (years) | Sample size | R<sup>2</sup> | MAE (age range) |
| --- | --- | --- | --- | --- | --- |
| **Here** | **Resting EEG** | **0 - 3** | **938** | **0.82** | **0.19 (0 - 2 years)**<br>**0.25 (0 - 3 years)** |
| [14] | sMRI | 0 - 2 | 76 | 0.79 | 0.15 (0 - 2 years) |
| [15] | sMRI | 0 - 5 | 220 | 0.97 | 0.19 (0 - 5 years) |
| [16] | sMRI | 0 - 3 | 658 | 0.85 | 0.19 (0 - 3 years) |
| [12] | Resting M/EEG | 0 - 95 | 2540 | 0.74 | > 6.48 (0 - 95 years) |
| [11] | Sleep EEG | 0 - 18 | 1056 | 0.96 | ~ 0.25 (0 - 2 years) |

**Table 3.** Sample Characteristics.

|  | Combined<br>Studies<br>N = 467 | Healthy<br>Baby<br>N = 13 | Infant<br>Sibling<br>N = 53 | Emotion<br>Project<br>N = 317 | Infant<br>Screening<br>N = 84 |
| --- | --- | --- | --- | --- | --- |
| **Sex, % Female (n)** | 49.0 (229) | 69.2 (9) | 52.8 (28) | 48.3 (153) | 46.4 (39) |
| **Ethnicity, % (n)** |  |  |  |  |  |
| Hispanic | 8.8 (41) | 23.1 (3) | 1.9 (1) | 11.0 (35) | 2.4 (2) |
| Non-Hispanic | 89.9 (420) | 76.9 (10) | 96.2 (51) | 87.7 (278) | 96.4 (81) |
| Not answered | 1.3 (6) | 0.0 (0) | 1.9 (1) | 1.3 (4) | 1.2 (1) |
| **Race, % (n)** |  |  |  |  |  |
| White | 77.9 (364) | 15.4 (2) | 83.0 (44) | 79.5 (252) | 78.6 (66) |
| Black/<br>African-American | 3.2 (15) | 46.2 (6) | 0.0 (0) | 2.2 (7) | 2.4 (2) |
| Asian | 4.1 (19) | 0.0 (0) | 3.8 (2) | 3.8 (12) | 6.0 (5) |
| More than one race | 12.8 (60) | 23.1 (3) | 11.3 (6) | 12.9 (41) | 11.9 (10) |
| Other | 0.6 (3) | 7.7 (1) | 0.0 (0) | 0.6 (2) | 0.0 (0) |
| Not answered | 1.3 (6) | 7.7 (1) | 1.9 (1) | 0.9 (3) | 1.2 (1) |
| **Family Income, % (n)** |  |  |  |  |  |
| <\$35,000 | 3.6 (17) | 23.1 (3) | 5.7 (3) | 2.2 (7) | 4.8 (4) |
| \$35,000 - \$75,000 | 10.7 (50) | 15.4 (2) | 7.5 (4) | 13.2 (42) | 2.4 (2) |
| >\$75,000 | 76.7 (358) | 38.5 (5) | 69.8 (37) | 76.0 (241) | 89.3 (75) |
| Not answered/<br>Don't know | 9.0 (42) | 23.1 (3) | 17.0 (9) | 8.5 (27) | 3.6 (3) |
| **Participant EEG data included in analysis,**<br>**n (% with longitudinal data)** |  |  |  |  |  |
| 50-140 days | 52 (92) | 13 (100) | 8 (100) | - | 31 (87) |
| 140-165 days | 101 (29) | - | - | 101 (29) | - |
| 165-250 days | 161 (49) | 12 (100) | 36 (100) | 113 (27) | - |
| 250-350 days | 52 (100) | 10 (100) | 42 (100) | - | - |
| 350-500 days | 224 (67) | 10 (100) | 46 (100) | 93 (22) | 75 (97) |
| 500-700 days | 86 (99) | - | 28 (100) | - | 58 (98) |
| 700-1000 days | 94 (99) | 10 (100) | 38 (100) | - | 46 (98) |
| 1000-1150 days | 168 (93) | 6 (100) | 38 (97) | 90 (89) | 34 (97) |
| By participant,<br>mean # EEGs (SD) | 2.0 (1.4) | 4.7 (1.2) | 4.5 (1.5) | 1.3 (0.4) | 2.9 (1.0) |

There are two notable findings related to our prediction error. First, we observed significant heteroskedasticity with increasing variance in predicted error with age. Brain volume dramatically increases in the first two years of life and then gradually slows down [27]; maximum synaptic density in middle frontal gyrus is reached at age 15 months, followed by synaptic pruning [3]. The fast changes in power spectra observed in infants match with these early, dynamic changes in the brain; once brain networks are more established in the second and third year, the power spectra becomes relatively stable [28]. This may explain the increased error variance at older ages. Second, as is commonly reported in brain age studies based on MRI data [17, 29, 30], we observed an “age bias”, characterized by the overestimation of age in younger children and underestimation in older children (Figure 2D). Age bias in predictive models is thought to be the direct result of fitting a regression model by minimizing the sum of the squares [31] and some studies have attempted to account for this through several bias correction methods performed after model estimation [17, 29, 30]. However, there are concerns that such correction may artificially inflate certain performance metrics such as the R^2^ values [31]. Thus, we have chosen to present uncorrected data, ensuring our findings are directly comparable to existing literature.

In this study, we also demonstrated a computational framework to estimate joint feature importance within clusters across highly correlated features. Analyzing feature importance quantifies the contribution of specific power measures on brain age prediction and may help understand the developmental trajectory of brain functions.

Using this method, important contributors to age prediction were identified including aperiodic offset and periodic power features in the low/high alpha, low beta, and theta band (Figure 3D and 3E). Developmentally, these features exhibit an approximately linear or monotonic relationship with age, which may have facilitated their feature importance in prediction models. Specifically, longitudinal modeling using generalized additive mixed models (GAMMs) of an overlapping data set recently demonstrated that aperiodic offset increases in the first year and then stays relatively stable between 1 – 3 years of age [28]. Aperiodic activity is hypothesized to be associated with strong broad band neural firing and the early increase is consistent with dramatic increases in gray matter volume and synaptogenesis observed in the first year [27, 32]. Theta and alpha oscillations have been implicated in thalamocortical circuitry development, and theta and alpha power also have more linear relationships with age [28, 33]. Alpha peak frequency shows age-dependent changes with peak frequency in the theta range in infancy, increasing to the alpha range by toddlerhood. This leads to a linear decrease in theta power from 1 – 3 years and a steady increase in high alpha power across the infant/toddler period. Linear increases are also observed in low beta band power across the first 3 years. In addition, a peak in the low beta range begins to emerge after the first year and is captured in the PC2 beta component included in the C1 cluster. These changes in beta power may represent the emergence of network connections between thalamic nuclei and cortical layers [33]. In summary, the identified importance of aperiodic offset and periodic power in theta, alpha, and beta bands may reflect their critical role in early brain development.

Our findings also support the use of EEG age prediction in early identification of neurodevelopmental delays. Age-predictions were significantly lower for two neurodevelopmental disorders associated with brain alterations and neurodevelopmental delay. The concept of a brain age gap — the difference between predicted and actual age — has been most thoroughly studied in brain degeneration [7, 13, 34, 35], brain injuries [36, 37], and mental disorders [38–40]. To date, we identified only three studies assessing individuals with neurodevelopmental disorders related to brain age gaps. Wang et al. [9] developed a prediction model using both structural and functional MRI data and reported significantly lower brain age gap in an ASD group (5 – 23 years) compared to a non-autistic group. They also observed associations between lower brain age gaps with lower ADOS scores within the ASD group. Tunc et al. [10] predicted brain age in individuals with ASD (6 – 25 years) using anatomical and diffusion metrics computed from MRI data and similarly observed that lower brain age was associated with higher ADOS severity scores. Iyer et al. [11] predicted age from sleep EEG and observed significantly lower brain age gap in children with DS (1 – 9 years) compared to the TD group.

Aligned with findings in prior literature, our EEG-based model was capable of generating distinguishable age predictions between ASD and TD groups at 3 years of age, and between DS and TD groups as early as 2 years of age (Figure 4A and C). We observed a negative and significantly lower brain-age gap for children with ASD (in the 1000 – 1150 days age bin) and DS compared to TD peers (Figure 4B and D). This preliminary data suggests that EEG brain gap measures could be clinically useful in identifying children with neurodevelopmental delays. Ideally such a measure would identify children before delays are present to facilitate earlier intervention. This will benefit children with disorders such as Fragile X syndrome and Tuberous Sclerosis Complex, where there are no obvious dysmorphisms at birth and diagnoses are often delayed. Future research with larger, diversified samples of both typically developing infants and infants with later identified delays are needed to build such clinical tools with sufficient accuracy.

It is important to clarify that the MLP model in this study was specifically developed to predict chronological age, not to identify individuals who might later be diagnosed with ASD or have DS. This distinction is crucial for interpreting our findings, particularly the group-level differences in brain age gap. While we observed significant negative brain age gaps in ASD and DS groups, this does not mean that a negative brain age gap is exclusively linked to ASD/DS, and it is not intended for diagnosing or screening for any neurodevelopmental disorder in particular.

The contribution of this study is subject to two major limitations. First, though we built our models using one of the largest developmental EEG data for our age range of interest, it is still not as large as the samples used in MRI-based age predictions [6, 13, 16, 41], and may even be considered small in the field of machine learning. Additionally, the majority of our data was collected from children who are disproportionately White and from families with a high socioeconomic status, lacking the diversity necessary to represent children of all backgrounds. Second, we acknowledge that more than one processing pipeline is available for developmental EEG [42–44], and the choice of pipeline and parameters may likely affect the output EEG, and thus the brain age prediction. Given it is impractical to evaluate our models across every processing configuration, we opted for one that we consider to be broadly reasonable. We are making our processing scripts, the parameters used, and the model training and testing scripts publicly available online, and welcome other researchers to test our method with their own data.

## 4 Conclusion

This study demonstrates the feasibility and accuracy of predicting chronological age from resting-state EEG features in infants and toddlers using a computational model. By analyzing data from a large cohort of typically developing children 2 to 38 months old, we identified significant EEG feature clusters that contribute most to age prediction. Moreover, the model’s ability to identify negative brain age gaps in children with autism spectrum disorder and Down syndrome suggests its potential as a tool for early detection of neurodevelopmental disorders. These findings support the clinical utility of EEG-based brain age prediction in monitoring and identifying early signs of developmental delays, enabling timely interventions.

## 5 Methods

### 5.1 Studies and Participants

EEGs for this paper were collected in a single lab at Boston Children’s Hospital as part of four different longitudinal studies. Sample numbers and demographics are shown in Table 3. Each study is described below.

Emotion Project (IRB-P00002876) was a cohort/longitudinal study with infants enrolled at either 5, 7, or 12 months of age and then followed through school age. EEG for enrolled infants was collected at age of enrollment and again at 3 years of age. While no developmental assessments were performed for this study, parents completed questionnaires regarding child development, interventions received, and diagnoses, including ASD diagnoses. Healthy Baby (IRB-P00019083) was a longitudinal study enrolling infants at 2 months of age with developmental assessments (Mullen Scales of Early Learning – MSEL [45]) and EEG collected at 2, 6, 9, 12, 24, and 36 months.

Infant Sibling Project and Infant Screening Project (IRB-X06-08-0374, IRB-P00018377) enrolled infants with and without a first degree family history of ASD (most often a full-sibling with autism) as early as 3-months of age. The Infant Screening Project also included infants with elevated social communication concerns at 12 months of age, and were excluded from this analysis. EEG and MSEL were administered at 3, 12, 18, 24, and 36 months across both studies, as well as 6 and 9 months of the Infant Sibling Project. Infants were assessed for ASD using the Autism Diagnostic Observation Schedule [46] or during the COVID pandemic some participants completed remote evaluations that included the Brief Observation of Symptoms of Autism [47], parent-child interaction, the Autism Screening Interview [48] and Vineland Adaptive Behavior Scales-Third Edition [49]. For toddlers meeting criteria on the ADOS or BOSA, a licensed clinical psychologist reviewed assessments and provided their best clinical judgement for ASD diagnosis based on criteria from the Diagnostic Statistical Manual of Mental Disorders, Fifth Edition [50].

All infants in the typically developing (TD) group were born with a minimal gestational age of 36 weeks, had no history of prenatal or postnatal medical or neurological disorders, and no known genetic disorders. Our age prediction model development was limited to participants without first degree family history of autism or known developmental delay or autism diagnosis. Exclusion based on developmental delays was determined as follows. Children with MSEL T-scores *<*40 on expressive language, receptive language, visual reception, or fine motor subscales at either 24 or 36 months were excluded. For children in the Emotion project where MSEL was not performed, children were excluded if parents reported a community diagnoses of speech or motor delay or if children were receiving Early Intervention services, or speech, occupational, or physical therapy services.

The ASD cohort (n = 85, 21 females; 246 EEG recordings, 15.8 *±* 9.6 months, range 2.1 – 37.7 months) included children who received a research-based clinical judgement of autism from Infant Sibling and Infant Screening Projects (n = 70, 214 EEG recordings), or children whose parents reported a community-based diagnosis (n = 15, 32 recordings) during follow up research visits.

The Down syndrome (DS) cohort (n = 21, 12 females; 21 EEG recordings, average age 26.4 *±* 6.0 months, range 13.9 – 36.8 months) included in this study was recruited as part of two separate studies collected in the same lab from 2019 – 2022 (IRB-P00018377, IRB-P00025806). DS participants all had Trisomy 21, a minimum gestational age of 36 weeks, and their families spoke primarily English at home (*>*50%). Children with known neurological disorders (e.g. intraventricular hemorrhage, seizure disorder) were excluded.

### 5.2 EEG collection and processing

Resting-state (non-task related) EEG data was collected using similar protocols across studies. The infant was seated with their caregiver in a dimly lit, sound attenuated room with a faraday cage to reduce noise. For the Emotion, Healthy Baby, Infant Screening Project studies a video of either infant toys or abstract moving objects were shown. For the Infant Sibling Project, a research assistant presented bubbles or showed toys to the infant in order to help the infant remain calm. Emotion, Healthy Baby, and Infant Screening Project studies used a 128-channel Hydrocel Geodesic Sensor Nets (Electrical Geodesics, Inc., Eugene, OR) connected to a NetAmps 300 amplifier (Electrical Geodesic Inc.) to collect continues EEG recordings with a 500Hz sampling rate. The Infant Sibling Project included recordings from either 64-channel Geodesic Sensor (*<* 10% of data) or a 128-channel Hydrocel Geodesic Sensor Nets (Electrical Geodesics, Inc., Eugene, OR), connected to either a NetAmps 200 or 300 amplifier (Electrical Geodesic Inc.) and sampled at either 250 or 500Hz. Data was referenced to the single vertex electrode, Cz, and electrooculographic electrodes were removed to improve infant comfort.

EEG raw files were collected in Netstation (Electrical Geodesics, Inc) and exported to MATLAB for preprocessing with the Batch Automated Processing Platform (BEAPP [51]) using the integrated Harvard Automated Preprocessing Pipeline for EEG (HAPPE [52]). A 1Hz high-pass and 100Hz low-pass filter were applied, and EEG data sampled at 500Hz were resampled to 250Hz. HAPPE artifact removal included 60Hz line noise removal, bad channel rejection, and artifact removal using first wavelet thresholding, followed by independent component analysis and Multiple Artifact Rejection Algorithm (MARA [53]). The following channels, in addition to the 10-20 electrodes, were used for MARA: 64-channel net – 16, 9, 8, 3, 58, 57, 21, 25, 18, 30, 43, 50, 53, 32, 33, 38, 41, 45; and 128-channel net – 28, 19, 4, 117, 13, 112, 41, 47, 37, 55, 87, 103, 98, 65, 67, 77, 90, 75, evenly covering all brain regions of interest. Next channels removed during bad channel rejection were interpolated, data were re-referenced to the average reference, detrended to the signal mean and segmented into 2-second segments. Segments were then further evaluated for and rejected for retained artifact using HAPPE’s amplitude and joint probability criteria.

EEG rejection criteria: HAPPE quality metrics were used to reject EEG recordings from subsequent analysis. EEG were rejected if they had fewer than 20 segments (40 seconds of total EEG), *<* 80% good channels, *>* 80% of independent components rejected, *>* 0.3 mean artifact probability of components kept, and *<* 25% percent variance retained.

### 5.3 Feature extraction

#### 5.3.1 Spectral parameterization

Using the BEAPP Power Spectral Density (PSD) module, the PSD at each electrode, for each 2-second segment, was calculated using a multitaper spectral analysis with three orthogonal tapers. For each electrode the PSD was then averaged across 2-second segments, and then averaged across electrodes in defined left frontal, right frontal, central, left temporal right temporal, and posterior regions of interest (Supplementary Figure 2).

The resulting PSD was then parametrized into aperiodic and periodic components using SpecParam v1.0.0 (ref; https://github.com/fooof-tools/fooof; in Python v3.6.8). SpecParam was modified for use in this age range — reason for and detailed description of modifications, along with modified code are discussed in [28]. SpecParam model parameters were: fixed mode, peak width limits set to [0.5, 18.0], max n peaks = 7, and peak threshold = 2. Detail evaluation of model fit across age ranges are also presented in [28].

Features extracted for age prediction include aperiodic parameters (offset and slope), periodic power across canonical frequency bands, and peak/amplitude characteristics of periodic peaks. Periodic power across canonical frequency bands [theta (4 – 6Hz), low alpha (6 – 9Hz), high alpha (9 – 12Hz), low beta (12 – 20Hz), high beta (20 – 30Hz), and gamma (30 – 45Hz)] was calculated. Peak characteristics were determined after the periodic spectrum was smoothed using a savgol filter (scipy.signal.savgoal filter, window length = 101, polyorder = 8). Kurtosis and skewness, characterizing the shape of power distributions, were calculated from the broad alpha range (4 – 12 Hz) and broad beta range (14 – 45 Hz) using scipy functions.

#### 5.3.2 Principal component analysis

We observed multiple shifts in periodic peaks with respect to age within the broad alpha range (4 – 12 Hz) and broad beta range (14 – 45 Hz) (Figure 1). Detailed characterization of these peak shifts utilizing an overlapping dataset are described in [28]. To capture this change across canonical frequency bands, we performed a principal component analysis (PCA) on the periodic power spectrum within each of these two broad ranges, identifying intrinsic functions that make up the spectrum. The top three PCs in each frequency range explain over 90% of the variance in the data and have distinctive peaks (Figure 1). The weight of each PC, quantifying the representation of each intrinsic function in data, was used as features in our age prediction models. This PCA step added six features (i.e., the weights of three PCs in each of the two frequency ranges) for each ROI, resulting in a total feature vector length of 72 (48 from spectral parameterization and 24 from PCA).

### 5.4 Regression models

We used Scikit-learn [54] to train and test four regression models for predicting chronological age using the extracted features. Prior to training, we preprocessed and transformed the data to enhance model performance. Each EEG feature was individually scaled to [0, 1] using a min-max scaler to avoid potential bias toward features with higher magnitude values and improve gradient descent optimization. This could also improve the optimization process by making the flow of gradient descent smoother. We then conducted recursive feature elimination (RFE) with a linear support vector regression kernel to remove features with low importance, resulting in approximately 60 retained features in each training iteration. For the dependent variable, we applied the Yeo-Johnson transformation to the chronological age to achieve a more Gaussian-like distribution, which is particularly important since we observed heteroscedasticity in the model prediction, as shown in Figure 2D.

#### 5.4.1 Lasso regression

We conducted experiments with elastic-net linear regression using varying ratios (0 to 1) of lasso and ridge regression. Our highest prediction accuracy was achieved with pure lasso. For L1 regularization, we set *α* to 0.001.

#### 5.4.2 Random Forest

We used 100 estimators (trees) with a maximum depth of 10. The maximum number of features in searching for the best split was set as the square root of feature length. Bootstrap was implemented when trees are being formed.

#### 5.4.3 XGBoost

XGBoost (Extreme Gradient Boosting,) is a gradient boosting decision tree framework. To prevent overfitting and ensure the generalizability of the learned model to new data, we employed a conservative parameter set for this algorithm. Specifically, we set a moderate maximum tree depth of 4, a low learning rate of 0.1, and a high *α* value of 0.1 for L1 regularization.

#### 5.4.4 Multilayer perceptron

A multilayer perceptron (MLP) is a type of feed-forward artificial neural network where all layers of neurons are fully connected and each neuron has an activation function to introduce nonlinearity to the model. In our experimentation with MLP models, we explored various architectures and hyperparameters, including the number of layers, the number of neurons in each layer, and levels of regularization. Our best performing model consisted of five hidden layers with 120, 120, 80, 60, and 40 neurons, respectively, with the rectified linear unit function serving as the activation function 2B. We set the *α* value for L2 regularization to 0.01, the batch size to 20, and gradually decreased the learning rate as a function of iteration number. Our findings indicate that MLP models with similar layer and neuron configurations to our best model typically yield comparable results.

### 5.5 Repeated cross-validation

We employed a 5-fold cross-validation to assess the performance of our models. To ensure that each fold had an equal number of samples across different age ranges, we divided the data into eight age bins, namely 0–140 days, 140–165 days, 165–250 days, 250–350 days, 350– 500 days, 500–700 days, 700–1000 days, and 1000-1150 days, based on the sample distribution (Table 3). For each iteration, we pseudo-randomly assigned samples into five equal folds and ensured that the distribution of samples in each age range was balanced across folds. We then trained the model on four folds and tested it on the left-out fold. We prevented information leakage by conducting PCA and all data preprocessing steps, including scaling, recursive feature elimination (RFE), and Yeo-Johnson transformation, only on the training set. We then applied the fitted transformers on both the training and test sets. The model’s performance was evaluated using the coefficient of determination (R^2^) value and the mean absolute error (MAE) of the test samples across all five iterations within the same train-test split. To reduce the influence of random data splits, we repeated this process ten times, each time with a different train-test split. We calculated the mean and standard deviation of the R^2^ and MAE values across the repetitions to verify the performance stability of our model.

### 5.6 Bagging cross-validated models

To further reduce the variance in predictions caused by the randomness in data splitting and improve model performance, we implemented the *bagging* technique, also known as *bootstrap aggregating*. We aggregated the models trained over ten repetitions of the train-test split. Following the repeated cross-validation described above, each sample in the dataset was tested ten times, each time using a slightly different model that was trained on a randomly drawn 80% of the total data (bootstrapping). However, it is possible that some of these models were suboptimal due to overfitting or underfitting, particularly with small sample sizes and excessive noise in data. Averaging the prediction results for a sample over multiple models (aggregating) can theoretically mitigate this problem, as such averaging can potentially remove undesirable characteristics of each model while synergizing their positive attributes [55]. We used the mean of ten predictions as the bagged prediction for a sample and calculated R^2^ and MAE using these bagged predictions.

### 5.7 Feature importance

In this study, a crucial question we aimed to address is which features in resting-state EEG drive age prediction. Though not detrimental to the overall prediction accuracy [56, 57], including highly correlated features in a regression model — especially features extracted from neighbouring electrodes or ROIs — would negatively impact the ability to accurately interpret the statistical importance of each independent variable (feature) [58, 59]. Here, we proposed a viable approach to circumvent this multicollinearity problem by evaluating feature importance at a cluster level using hierarchical clustering and Shapley Additive Explanations (SHAP) values (Figure 3A).

#### 5.7.1 Shapley values

The Shapley value employs concepts in game theory and fairly distributes both gains and costs to features working in coalition. Specifically, each feature of an observation is assigned with a Shapley value, which equals the amount that this feature steers the model away from a baseline prediction (the sample mean). Using the SHAP python package [60], we calculated SHAP values from each sample in the TD group via a 5-fold cross-validation, in which an MLP model was trained with 4 folds of data, and feature importance was calculated for the left-out fold (test fold). This process was repeated five times, each with a different test fold, so that each sample was tested once, and hence had one set of SHAP values.

#### 5.7.2 Feature clustering

We grouped all EEG features into clusters based on the Pearson’s correlation between each pair of EEG features and used one minus the absolute value of this correlation as a measure of distance between features. Then we applied the Ward hierarchical clustering to group features into clusters. By setting a reasonable distance threshold, we divided the 72 features into 11 clusters (Figure 3B).

#### 5.7.3 Cluster-SHAP

Employing its additivity property, we summed SHAP across features in each cluster for their joint SHAP score (Cluster-SHAP). We then compared Cluster-SHAP among feature clusters to evaluate their relative importance. Cluster importance can then be evaluated from the mean absolute value of Cluster-SHAP across samples and the correlation between Cluster-SHAP and actual age. In both metrics, a higher value indicates greater cluster importance [26].

### 5.8 Prediction on children with autism or Down syndrome

We further examined our MLP model’s sensitivity in identifying potential neurodevelopmental delays by testing it on data from the ASD and the DS groups. For the ASD group, we first age-matched each sample (n = 85, 246 EEG recordings) with one from the TD group (test set). Then, we retrained the MLP model with the remaining TD samples and applied this new model on the test set. Similarly, for the DS group, we age-matched each sample (n = 21, 21 EEG recordings) with one from the TD group, retrained the MLP model with the remaining TD samples, and compared prediction errors between groups.

### 5.9 Statistical analysis

The correlation between predicted age and actual age and between Cluster-SHAP and actual age was evaluated by Pearson’s r using the Scipy function. Group-level difference in age prediction error and EEG features was examined by a two-sample t-test.

## Supplementary information

Supplementary information for this paper includes two figures.

## Acknowledgements.

We thank all the children and families who generously participated in this research. We thank all the research staff involved in participant recruitment, data collection, and database administration. We thank Dr. Helen Tager-Flusberg, Dr. Michelle Bosquet Enlow and Dr. Nicole Baumer for acquiring funding and helping to oversee projects that contributed data to this study.

## Author contribution

Winko W. An: Methodology, Software, Formal analysis, Writing Original Draft, Visualization. Aprotim C. Bhowmik: Portions of Formal Analysis, Writing – Review and Editing. Charles A. Nelson: Investigation, Resources, Writing – Review and Editing, Supervision, Project administration, Funding acquisition. Carol L. Wilkinson: Conceptualization, Investigation, Resources, Data Collection, Data Curation, Writing – Portions of Original Draft, Review and Editing, Supervision, Project administration.

## Data availability

Consents obtained from human participants prohibit sharing of deidentified individual data without data use agreement in place. Please contact the corresponding author with reasonable data requests.

## Code availability

Code used for EEG processing and analyses used in this paper can be found on the Open Science Framework (https://osf.io/u3gp4).

## Ethics approval and consent to participate

All studies that contributed data to this work were reviewed and approved by IRB at Boston Children’s Hospital (IRB-P0000287, IRB-P0001908, IRB-X06-08-0374, IRB-P0001837, IRB-P00018377, IRB-P00025806). Written consent was obtained prior to subject participation.

