## Supplemental figures 1 and 2 for "Prediction of chronological age from resting-state EEG power in the first three years of life"

### Supplementary materials

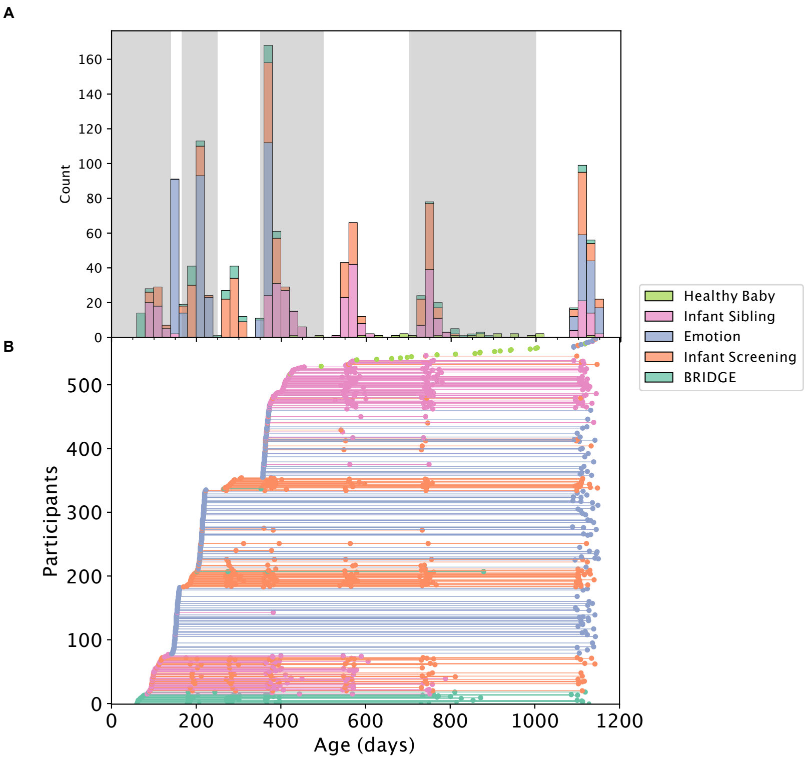


**Fig. 1** A: Histogram of participants with typical development or autism spectrum disorder across age. Shaded areas represent the age bins. B: Subject contribution of data across age. Four longitudinal studies from which data were pulled are coded in different colors.

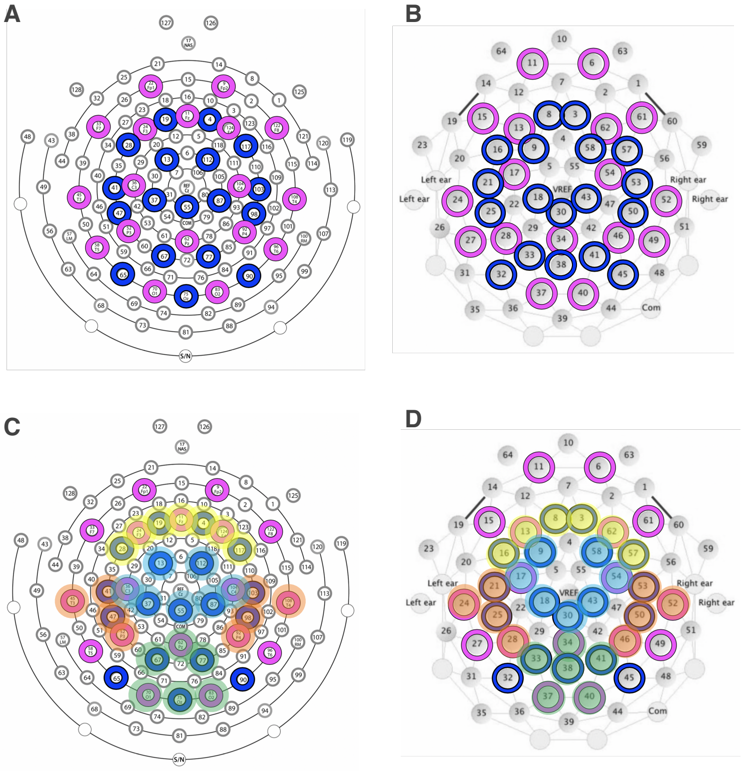


**Fig. 2** Electrode layout. A: 128-channel Hydrocel Geodesic Sensor Net. B: 64-channel Geodesic Sensor Net. Pink circles denote 10-20 electrodes, and blue circles denote the additional electrodes included in ICA and MARA steps of pre-processing. C and D: Electrodes averaged for frontal (yellow), central (blue), temporal (orange), and posterior (green) regions of interest.
